# Durability of second-line anti-retroviral therapy in a resource-limited setting: an 11-year analytical cohort

**DOI:** 10.1101/2022.03.15.22272370

**Authors:** I. Lumu, J. Musaazi, B Castelnuovo

## Abstract

**Background:** It is projected that up to 19·6% of patients on ART in Sub-Saharan Africa will need second-line treatment by 2030, but the durability of such therapy remains unclear. This study investigated the durability of second-line ART and the factors associated with the viral rebound among patients on second-line ART in Uganda.

**Methods:** A retrospective dynamic cohort of adults initiated on second-line ART after confirmed virological failure to first-line ART. Patients that had taken second-line for ≥6 months between 2007 and 2017 were included. Patients were followed until they experienced a viral rebound (Viral load ≥200copies/ml). Cumulative probability of viral rebounds and factors associated with viral rebound were determined using Kaplan-Meier methods and Cox proportional hazard models, respectively.

**Findings:** 1101 participants were enrolled. At base-line, 64% were female, the median age was 37 years (IQR 31-43), median duration on first-line ART was 44months (IQR 27-67), and the median CD4 and viral load were 128 cells/ul (IQR 58-244) and 45978 copies/ml (IQR 13827-139583), respectively. During the 4757·21 person-years, the incidence density of viral rebound was 74·62 (95% CI 67·25- 82·80) per 1000 person-years. The probability of a viral round at 5 and 10 years was 0·29 (95% C: I 0·26 -0·32) and 0·623 (95%CI:0·55 -0·69), respectively. The median survival without experiencing a viral rebound was 8·7 years. Young adults (18-24) years (aHR 2·31 95 CI 1·25-4·27), high switch viral load ≥100,000copies/ml (aHR 1·53 95 CI 1·23- 1·91) and ATV/r based second-line (aHR1·53 95 CI 1·18-2·00) were associated with an increased risk viral rebound.

**Interpretation:** Second-line regimens are fairly durable for eight years followed a rapid increase in the incidence of rebounds. A high viral load at switch, ATV/r based second-line, and young adulthood are risk factors associated with a viral rebound, which underscores the need for differentiated care services.

## INTRODUCTION AND BACKGROUND

The introduction of Antiretroviral therapy (ART) and expansion of the ART program as a prevention strategy has been the most effective intervention for reducing the incidence of HIV in the population.^1^ To this end, Joint United Nations Programme on HIV and AIDS (UNAIDS), has advocated for the further expansion of the ART program as a means of elimination of AIDS suggesting that the 95-95-95 will result in a reduction in total population HIV viral load, and thus halt the spread of HIV and stop the AIDS epidemic. ^2,3^

With the current expansion of the ART program, about 23 million people are on ART globally, and about 66% of these are in sub-Saharan Africa.^4^ Among the patients on treatment in developing countries, 5.9% of them are on second-line treatment after failing the initial therapy,^5,6^ but this proportion is projected to increase with the growing acquired resistance to first-line,^7^ and the increased viral load monitoring.^8^ Mathematical modelling projects that about 20 million people in Sub-Saharan Africa (SSA) would need ART by 2030, and 19.6 percent of these would be on second-line treatment^8^. Second-line therapy is expensive and costs about 2·5 times more than first-line, which means that SSA countries must budget for their second-line ART needs.^8,9^ Meanwhile, some studies show that up to 38% of patients starting second-line therapies in SSA fail treatment within three years,^10^ and are frequently maintained on the same regimen since third-line drugs are not widely available in most public health programs^3^. These patients remain at risk of experiencing an AIDS event and pose a risk of transmission of HIV and thus undermining global public health efforts to halt the HIV epidemic.^3,13,14^ It is essential, therefore, that patients switched maximise the benefit of second-line treatments.

Although clinical trials show that the World Health Organisation (WHO) recommended second-line treatments are efficacious,^15-16^ these trials are usually short and unable to predict the long-term durability of these regimens.^17^ Therefore, this retrospective cohort aimed to determine the durability of second-line treatment and factors associated with viral rebound in a resource-limited high-burden setting.

## METHODOLOGY

### Study design and setting

This was a retrospective dynamic cohort based on electronic medical records including data recorded between 1st January 2007 and 31^st^ December 2017. The study was conducted at the Infectious Disease Institute(IDI), Makerere University in Kampala, Uganda. The institute is a centre of excellence in HIV care and receives referrals from other clinics around the country. Currently, the HIV clinic has about 8000 active patients receiving long term care. About 1450 (20%) of active patients are on second-line treatment. However, all patients that had taken second-line during the study period and met the exclusion criteria below were considered for analysis irrespective of their current vital status.

On each visit, an intranet-based system is used to enter a patient’s data in real-time. The data collected during the visit relates to HIV disease monitoring and ART adherence. The centre follows the WHO guidelines on treatment care and prevention. However, IDI being is a research setting, more frequent viral load monitoring may be done. To ensure completeness of clinical notes and compliance with the guideline, the clinic has an active quality control department.

### Definition and procedures

In this study, viral rebound was defined as a single HIV-RNA viral load ≥200 copies per ml at least six months after switching to second-line therapy. Durability was defined as the time a patient spends on second-line ART without experiencing viral rebound after achieving viral suppression, that is, the duration of a sustained viral suppression after at least six months on second-line therapy.

The study included adults that were aged ≥18 years at the time of starting second-line, and who had been switched to second-line after confirmed virological failure of first-line treatment and had stayed on the new regimen treatment for ≥ 6 months. The six-month period was a proxy to ensure that only individuals that had a chance of achieving viral suppression entered the analysis. The study excluded patients without a viral load to confirm treatment failure at the time of switch to second-line, patients without a clear history of first-line treatment, and patients that did not have follow-up viral loads during treatment (Figure 1).

**Figure 1:**
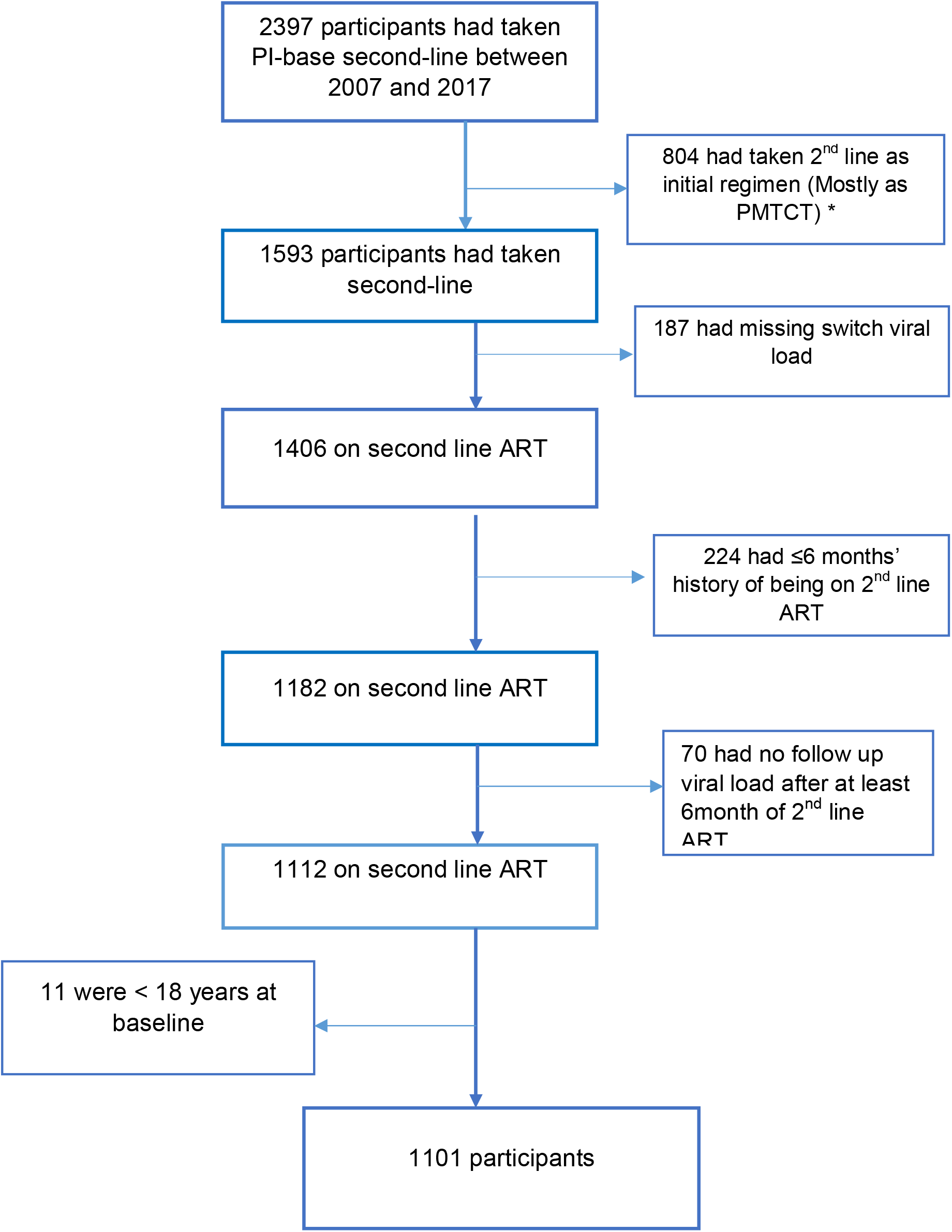
Diagram illustrating how participates were enrolled into study. *PMTCT- Prevention of Mother to Child Transmission of HIV

Patients were followed-up from the date of switching to second-line until they experience a viral rebound viral load, died, got lost to follow-up, or transferred whichever event occurred first. For these events, censorship was done on the date of the last visit. For participants without the outcome of interest, data were administratively censored on the 31st of December 2017.

### Statistical Analysis

Statistical analysis was performed using STATA version 15·0. Baseline participants’ characteristics were described using frequency and percentages for categorical variable, median, and interquartile range (IQR) for continuous variables. We calculated the incident density of viral rebound per 1000 person-years during the eleven.^13^ The study compared the incidence of viral rebound with alternative definitions. In the sensitivity analysis the study defined viral rebound, as having a single HIV-RNA viral load ≥1000 copies/ml. Also, we modelled for virological failure defining it as having two consecutive viral loads≥ 200 copies/ml or ≥1000 copies/ml within 2 years from the first viral load.

Kaplan-Meier methods were used to determine the time to first viral rebound and the median survival without a viral rebound over the 11 years reflected the durability of second-line treatment. The cox-proportional hazard (Cox-PH) model was used to determine the patients’ factors independently associated with the first viral rebound. Patients’ factors explored included: age, sex, viral load, CD4 count, duration on first-line, Hepatitis B co-infection, TB co-infection, Nucleoside Reverse Transcriptase Inhibitor(NRTI) back backbone, None-Nucleoside Reverse Transcriptase Inhibitor(NNRTI), and anchor Protease Inhibitor (PI). The continuous variables were categorised into groups to reflect clinical or public health approaches to managing HIV/ AIDS. In the adjusted model, variables with p-value<0·2 at unadjusted analysis were selected into the adjusted model. Proportional hazard assumption for Cox-PH was checked using Schoenfeld residuals. The final model was checked for collinearity and effect modification between covariates. Statistical significance was set at 5% level. Sensitivity analysis using death and loss to follow-up as competing risks with viral rebound, was performed using Cumulative Incidence Function.

### Ethical considerations

Ethics approval for the study was obtained from the University of Liverpool Research Ethics Committee in the United Kingdom. In Uganda, permission to conduct the study was obtained from the Scientific Review Committee of the Infectious Diseases Institute. At this institute, approval for the analysis of routinely collected data and the waiver for informed were granted by the Institutional Review Board of Makerere University Faculty of Medicine (approval number: 120-2009) and the Research Ethics Committee of the Uganda National Council for Science and Technology (approval number: 45683) under the routine care protocol. This approval and waiver are reviewed annually by both the Institutional Review Board of Makerere University Faculty of Medicine and the Research Ethics Committee of the Uganda National Council for Science and Technology.

### Role of funding

No funding was obtained for this study. The corresponding author had full access to all the data and the final responsibility and decision to submit the results for publication.

## RESULTS

During this period (2007 and-2017), we identify 2397 patients that had taken PI-based second-line regimen. Eight hundred four (33·5%) had taken this regimen as initial therapy and 187(7·8%) had no viral loads at switch. Two hundred twenty-four (9·3%) had taken second-line for less than six months, 70(2·9%) patients did not have a follow viral load, and 11(0·5%) patients were less than 18 years at baseline. A total of 1101 were included in the analysis (Figure 1). In this cohort, the median age was 37 years (IQR 31-43) and 705(64%) of the patients were female. At baseline, the median viral load was 45978 (IQR 13827-139583) copies/ml with 68% of the participants having a viral load<100,000 copies/ml at switch. Only 956(87%) of patients had a CD4 count at switch with a median of 128 (IQR 58-244) cells/ul. At switch, 792(72%) of the patients were changed to TDF-based NRTI backbone and 561(51%) were given boosted LPV/r as the anchor PI. One hundred and eleven (10%) were diagnosed with TB during the 11-years period. Forty-two (4%) of the participants were co-infected with hepatitis B (Table 1).

**Table 1:** Demographic and clinical characteristics of the participants.

| Characteristics | Number of participants<br>N=1101(%) |
| --- | --- |
| **Age (years), median (IQR)** | **37( 31-43)** |
| **Categories, n (%)** |  |
| 18-24 | 68(6) |
| 25-34 | 359(33) |
| 35-44 | 444(40) |
| 44-55 | 175(16) |
| >55 | 55(5) |
| **Sex, n (%)** |  |
| Male | 396(36) |
| Female | 705(64) |
| **Viral load at switch copies/ml, median (IQR)** | **45978( 13827-139583)** |
| <1000 copies/ ml | 39(4) |
| 1000-<10,000 | 192(17) |
| 10,000-<100,000 | 519(47) |
| ≥ 100,000copies/ml | 351(32) |
| **CD4 at switch (cells/ul) median(IQR)** | **128(58-244)** |
| <200 | 644(58.5) |
| ≥200 | 312(28.3) |
| Unknown | 145(13.2) |
| **Duration of First-line(ART)(months) median (IQR)** | **44 ( 27-67)** |
| 0-36 | 429(39) |
| 48-60 | 317(29) |
| >60 | 355(32) |
| **NNRI Used in first line** |  |
| EFV-based | 388( 35) |
| NVP-based | 660( 60) |
| Others | 53( 5) |
| **Second-line((NRTI backbone)** |  |
| TDF-based | 792(72) |
| AZT-based | 98(9) |
| **Others** | 211(19) |
| **Second-line PI** |  |
| **ATV/r-based** | 540(49) |
| **LPV/r-based** | 561(51) |
| **Calendar Year of initiation of second line** |  |
| **2007-2010** | 256(23) |
| **2011-2014** | 525(47) |
| **2015-2017** | 320(29) |
| **CDC score at switch** |  |
| **Working** | 60(5.4) |
| **Bedridden or Ambulatory** | 665(60.4) |
| **Unknown** | 376(34.2) |
| **Hepatitis B co-infection** |  |
| **Negative** | 1059(96) |
| **Positive** | 42(4) |
| **TB co-infection** |  |
| **No** | 990(89.9) |
| **Yes** | 111(10.1) |
| **Adherence** |  |
| **Good** | 996(90.5) |
| **Fair** | 6(0.5) |
| **Poor** | 21(1.9) |
| **Unknown** | 68(6.2) |

At the closure of the database, 683 (62%) patients had not experienced a viral rebound 355 (32.2%) had a viral rebound. Sixteen (1·5%) patients were transferred out, 23 (2·1%) got lost to follow-up, and 24 (2·2%) had died before experiencing a viral rebound ≥200 copies/ml. Viral rebounds are described by patients’ characteristics in Supplementary Table 1. For the 1101 individuals, the total time at risk was 4757·21 person-years and the incidence density of a viral rebound (≥ 200copies/ml) was 74·6 2(95% CI 67·25- 82·80) per 1000person-years. The incidence of viral rebound at one year was 78·56(95% CI 63·51-97·17) 1000 person-years. At three, six, and nine years the rate was 67·21(95%CI 51·08-88·44) per 1000 person-years, 78·64(95%CI 53·93-114·68) 1000person years, 113·95 (95%CI 63·10-205·76) respectively (figure 2 and figure 1s).

**Figure 2:**
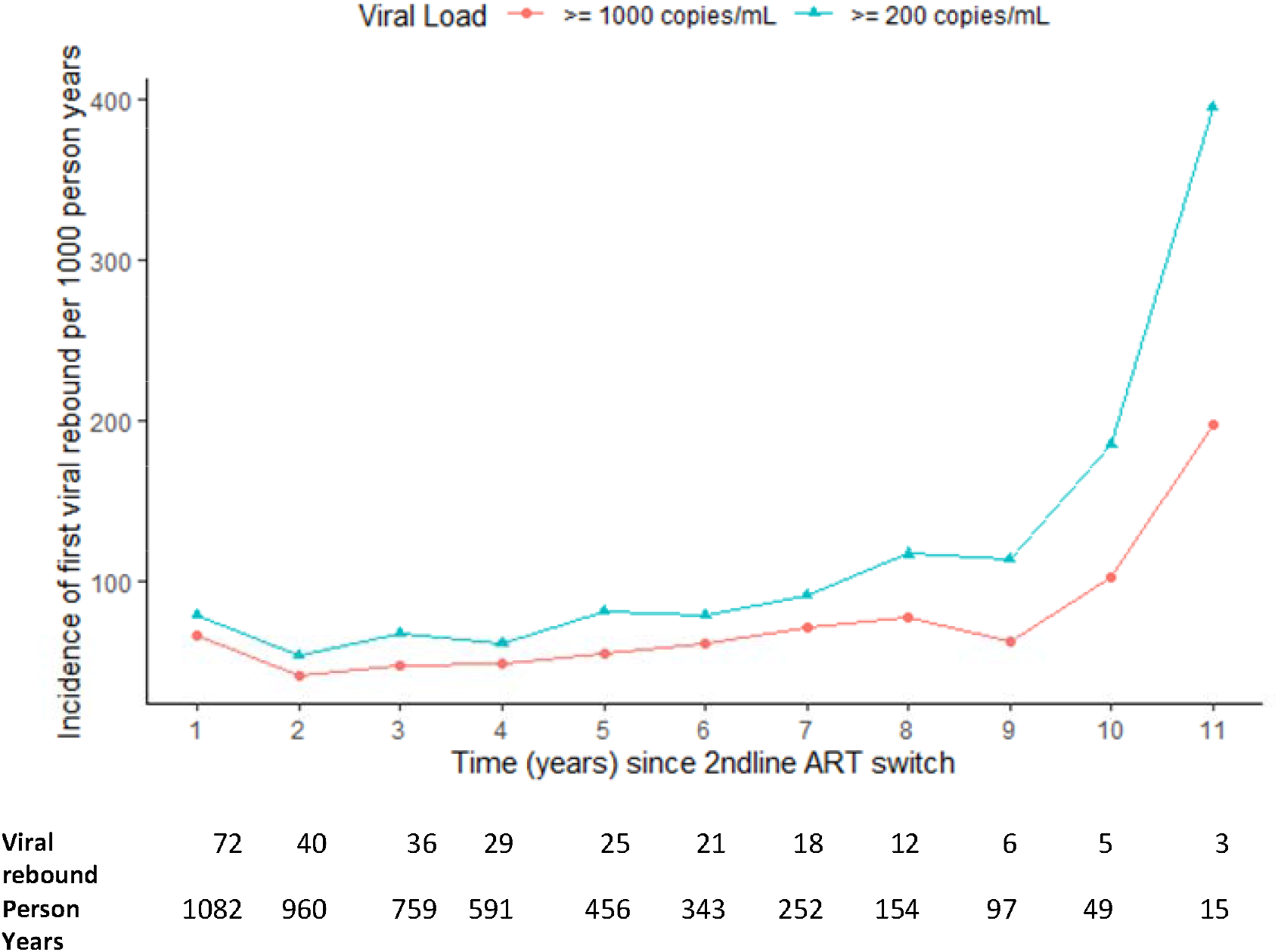
Incidence of viral rebound (≥ 200 copies/ml) after switching to secondline.

When we used a different cut of a viral rebound ≥ 1000 copies/ml, 267(24%) had a viral rebound and 834(76%) had not rebound. In patients with viral rebounds, I88 (52%) had two consecutive viral loads of ≥200copies/ml and 153 (57%) had two consecutive viral loads ≥1000 copies/ml. The incidence of viral load rebound was 56·13(95% CI 49·78-63·27) per 1000person-years when we considered a viral rebound at ≥1000 copies/ml at switch.

The median survival without a viral rebound after 4757·21 personal years was 8·7 years. At one year, the probability of rebound was 0·087 (95% CI: 0·06- 0·09). At two years, the cumulative probability of rebound was 0·13 (95% CI 0·11 - 0·15). At three years, the probability was 0·18(95% CI: 0·16- 0·21), and 0·29 (95% CI 0·26 -0·32) at 5 years. The cumulative probability of rebound at 10 years was 0·623 (95%CI:0·55 -0·69) (Figure 3a). The cumulative probability of viral rebound by switch viral load was 0·65(95% CI 0·56-0·73) and 0·62 (95% CI 0·52- 0·72) for a switch viral load≥100,000 copies/ml and <100,000 copies/ml, p-value <0.01 (figure 3b). When the median survival without a rebound was assessed using the ≥1000 copies/ml cut-off, rebound-free survival was 10·6 years an increase of two years. Similarly, the study assessed the effect of competing risks (death and loss to follow-up). The Kaplan Meir and Cumulative Incident Function were too close. This implies the impact of death and loss to follow up on incidence rate estimates is negligible. The study also separated explored the selection bias due to loss to fool-up assuming that all LTFU as a viral rebound and there was no difference in survival estimates.

**Figure 3a.**
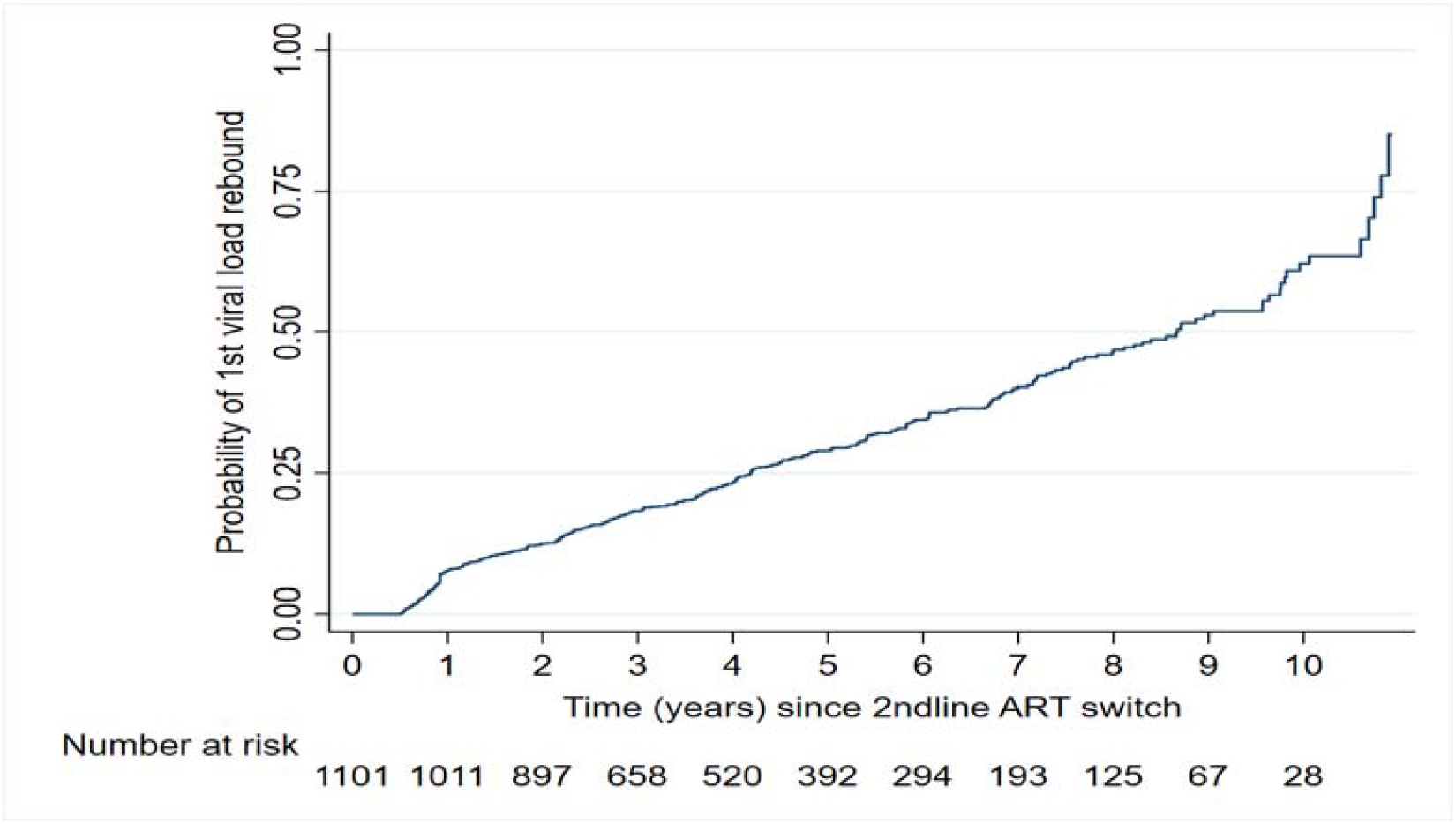
Kaplan-Meier plot for the probability of having first viral load rebound.

**Figure 3b:**
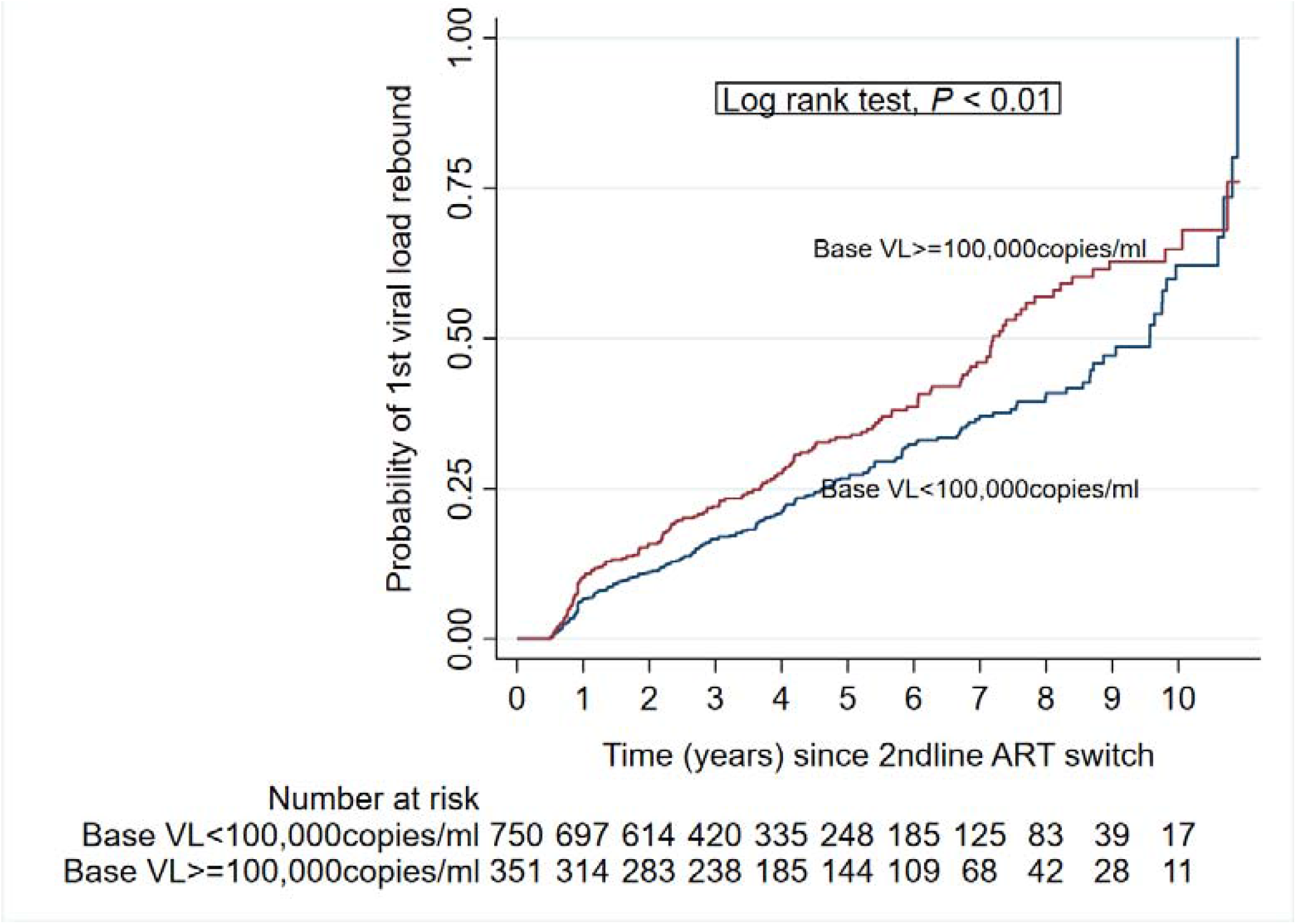
Kaplan-Meier plot for the probability of having first viral load rebound stratified by baseline viral load categories at 2ndline ART switch.

In the univalent analysis, being a young adult (18-24 years) compared to being older (>55 years) was associated with a viral rebound (uHR 2·59 1·41- 4·73, p= 0·002). Using EFV-based NNRTI in first-line (uHR 1·60 95%1·29- 1·99 p<0·001), a switch viral load of ≥100,000copies (uHR 1·36 95% CI1·10-1·68, p-value 0·005), using ATV/r anchor PI (uHR 2·22 95% CI 1·75- 2·82, p<0·001), and calendar year of initiation of second-line 2011-2014, 2015-2017 were associated increased viral rebound (Table 2). In multivariate analysis, these associations persisted (Table 2). A viral load ≥100,000 copies/ ml at time of switch (adjusted HR: 1·45, 95%CI: 1·20- 1·86, p-value< 0·001), calendar year of initiation of second-line, 2011-2014 (aHR: 1·51, 95%CI: 1·15-1·99, P=0·003), and 2015-2017 (aHR: 2·83, 95%CI: 1·26- 2·23, P-value<0·001) as compared to 2007-2010, history of EFV (aHR1·49 95 CI 1·18-1·86 p-value 0·001), boosted ATV/r (aHR:1·53 95 CI 1·18-2·00 p-value 0·002), and being 18-24 years (aHR: 2·31 95 CI 1·25-4·27 p-value=0·007) were all associated with viral rebound. In the sensitivity analysis, there was no difference in the estimated adjusted Hazard Ratio when a high viral load cut of ≥1000 copies/ml was used and the same factor remained associated with viral rebound but with increased Hard Ratios. Similarly, the sub-distribution hazard model found the same factors to be associated with viral rebound (Supplementary table 2).

**Table 2:** Cox-proportional hazard model for viral rebound and baseline factors associated.

| Variable | Viral rebound $\geq 200$ copies/ml<br>n=355(%) | Unadjusted (95%CI) | p-value | Adjusted HR (95%CI)† | p-value |
| --- | --- | --- | --- | --- | --- |
| **Age** |  |  |  |  |  |
| 18-24 | 35/68 (51) | 2.59 (1.41- 4.73) | 0.002 | 2.34 (1.26-4.32) | 0.007 |
| 25-34 | 114/359 (31) | 1.06 (0.62- 1.81) | 0.837 | 1.16 (0.67- 2.00) | 0.603 |
| 35-44 | 133/444 (30) | 0.97(0.57- 1.65) | 0.907 | 0.95(0.55 -1.62) | 0.841 |
| 44-55 | 58/175 (33) | 1.14 (0.65- 2.03) | 0.635 | 1.14 (0.64- 2.02) | 0.663 |
| >55 | 15/55 (27) | 1 |  | 1 |  |
| **Sex** |  |  |  |  |  |
| Male | 144/396 (34) | 1 |  | 1 |  |
| Female | 211/705 (30) | 0.88 (0.70-1.10) | 0.110 | 0.87 (0.69- 1.09) | 0.087 |
| **Viral load at switch**<br>**copies/ml** |  |  |  |  |  |
| <100,000 | 208/750 (28) | 1 |  | 1 |  |
| ≥100,000 | 147/351 (42) | 1.36 (1.10-1.68) | 0.005 | 1.52 (1.21- 1.90) | <0.001 |
| **CD4 at switch**<br>**(cells/ul)** |  |  |  |  |  |
| <200 | 212/644 (33) | 1 |  |  |  |
| ≥200 | 92/312 (29) | 1.15 (0.90-1.46) | 0.276 |  |  |
| **Duration of**<br>**First-**<br>**line(ART)** |  |  |  |  |  |
| 0-3 | 149/429 (35) | 1 |  |  |  |
| 4-5 | 106/317 (33) | 0.89 (0.69-1.14) | 0.351 |  |  |
| >5years | 100/355 (28) | 1.02 (0.79-1.32) | 0.869 |  |  |
| **First-line**<br>**NNRTI used** |  |  |  |  |  |
| EFV-based | 145/388(32) | 1.60(1.29-1.99 ) | <0.001 | 1.49(1.18-1.86) | 0.001 |
| NVP-based | 190/660( 29) | 1 |  |  |  |
| Others | 20/53( 38) | 1.59(1.00<br>2.52) | 0.049 | 1.66(1.03- 2.67 ) | 0.004 |
| **Second-line**<br>**NRTI** |  |  |  |  |  |
| TDF- based | 255/792 (32) | 1 |  | 1 |  |
| AZT-based | 29/98 (30) | 1.42 (0.96 -<br>2.09) | 0.079 | 0.83(0.55-1.26) | 0.393 |
| Others | 71/211 (34) | 1.03 (0.96 -2.09) | 0.079 | 1.18(0.88-1.57) | 0.259 |
| **PI used in**<br>**second-line** |  |  |  |  |  |
| LPV/r -based | 189/561 (32) | 1 |  |  |  |
| ATV/r-based | 166/540 (31) | 2.22 (1.75-2.82) | <0.001 | 1.71 (1.28-2.29) | <0.001 |
| **Calendar**<br>**Year of**<br>**initiation of**<br>**second line** |  |  |  |  |  |
| 2007-2010 | 95/256 (37) | 1 |  | 1 |  |
| 2011-2014 | 177/525 (33) | 3.09 (2.21-4.30) | <0.001 | 2.49 (1.75-3.56) | <0.001 |
| 2015-2017 | 83/320 (26) | 6.96 (4.60-10.54) | <0.001 | 5.53(3.52- 8.71) | <0.001 |
| **Hepatitis B**<br>**co-infection** |  |  |  |  |  |
| Negative | 342/1059 (32) | 1 |  |  |  |
| Positive | 13/49 (31) | 1.05 (0.60- 1.82) | 0.868 |  |  |
| **TB co-**<br>**infection** |  |  |  |  |  |
| No | 214/890 (32) | 1 |  |  |  |
| Yes | 41/111 (37) | 1.26 (0.91- 1.74) | 0.282 |  |  |

† Adjusted results were similar when death and lost to follow-up were analysed as competing risk events for viral rebound using sub-distribution hazard model.

## DISCUSSION

Among ART experience patients that are starting second-line treatment after virological failure of first-line therapy, viral rebound is common with an incidence of 7·5 per 100 person-years. The incidence of viral rebound did not differ much when a different cut-off was used implying that stringent measurement was effective at identifying more patients at risk of failure. Over time we observe a reduction in incidence from the time of switch to six years. After this period the incidence increases gradually to eight years followed by a rapid increase in viral rebounds. Moreover, between five and ten years the cumulative probability of viral rebound doubled from 29% to 62%. It appears the longer patients spend on second-line ART the higher the likelihood and incidence of a viral rebound. We think that this is probably due to complacency and fatigue after being on ART for a long time. Secondly, this could be due to the acquisition and accumulation of more drug mutations over time hence the increased resistance that eventually results in viral rebound. Nonetheless, the observed median rebound-free survival of eight years is reassuring and suffice to say that current treatment regimens used in this setting are fairly durable. However, after this time the probability and incidence of rebound increase rapidly although the confidence interval is wide due to reduced person-term in this dynamic cohort. Of note, this does not put a limit on the durability of ART in individual patients with favourable traits and a good routine of taking medication that may survive longer.^19^ The rebound free survival reported here is higher than that reported by some studies in high-income settings. ^20,21^ This is probably because of the difference in the type of second-line used in these studies.

The incidence of viral rebound was significantly higher in patients that had a viral load ≥100,000 copies/ml at switch. Similarly, we observed a significantly lower median rebound-free survival in this cohort compared to those with a viral load of <100,000 copies/ml at switch. In the multivariate analysis, these individuals were 52% more likely to experience a viral rebound. This may be because such patients with a high viral load at switch either have a high viral reservoir,^22^or are switched late after accumulating several drug mutations to the backbone regimen,^23,24,25^ and thus may be taking regimens with just one-two active components. Also, such people are likely to have suboptimal adherence and this behaviour continues after switching. Altogether these factors render them more susceptible to viral rebounds.

Individuals that were switched to boosted ATV(ATV/r) were 70% more likely to experience a viral rebound compared to those taking LPV/r. Although the WHO recommends both protease – inhibitors, ATV/r has a lower genetic barrier to resistance than LPV/r.^26^ Moreover, there is limited evidence to support the use of ATV/r in second-line regimens.^27^ Therefore, taking a drug with a lower genetic barrier will lead to rebound after a short while when patients have acquired and accumulated sufficient mutations. Similarly, the patients who had switched from EFV-based regimens had a 49 % risk of viral rebound compared to those taking Nevirapine. The reasons for this are not clear but we think they may be related to differences in adherence behaviour in this sub-groups of patients. However, both treatments have a low genetic barrier and cross-resistance,^26^ and are being rolled back due to high levels of primary resistance.^28^ On the other hand, we did not find an association between viral rebound and the type of NRTI backbone regimen. Currently, once patients fail on TDF based regimen as first-line they are switched to AZT-based second-line and vice versa despite the suspected cross-resistance among NRTIs. This finding is in agreement with previous clinical trials done in resource-limited settings.^15,16^ The EARNEST and NADIA trial suggest that NRTI backbones remained effective as second-line therapy even in patients with multiple mutations.^15,16,29^

We observed that later years of switch were associated with viral rebound, contrary to previous studies that showed that later years were associated with a lower rate of viral failure because of the introduction of newer durable regimens that are easy to administer.^21,30^ The year categories in this study were chosen to reflect changes in treatment guidelines by WHO and this finding may be attributed to the change in public health policy to repeat viral load every six months from twelve months which may have improved detection of viral rebounds.^31, 32^ However, our clinical experience indicates that patients initiating treatment in later years tend to be less adherent compared to previous years. Also, the study found that young adults have a two-fold increased risk of experiencing a viral rebound compared to older age groups. This finding is similar to observations of a previous study on second-line treatment outcomes.^33^ Older patients tend to be more adherent,^19^ stable socially and economically compared to young people, and thus, have better adherence and compliance with care.^34^

Since most resource-limited and high-burden settings use WHO PI-based second-line treatments, the findings offer insight into the durability of the current treatments, the expected number of viral rebounds in the clinic, and the need for third-line treatment. This study identifies a population of patients that need differentiated services, for example, young adults who should be provided with youth-friendly services and close monitoring. Moreover, we suggest that clinicians need to consider all the modifiable risk factors before choosing the appropriate second-line regimen. We recommend the provision of frequent clinical monitoring, enhanced adherence counselling, frequent viral load monitoring, measurement of drug levels, and drug resistance testing as part of the differentiated service to risk groups although this may be challenging as a public health approach. Also, we think that HIV patients should be monitored closely and reoffered special adherence sessions after six years on second-line ART.

Our findings have implications for HIV programs and clinics in resource-limited settings that are using WHO-recommended second-line ART. The observed durability in this study supports the continued use of these regimens since they are affordable.^3^ However, the increased risk of viral rebound associated with ATV/r calls for consideration of alternative single-dose regimens like Dolutegravir, but the evidence supporting its use after failure is still limited. The ongoing NADIA trial has shown that 90% of patients on DTG –based second-line achieve suppression by forty-eight weeks.^29^ These updates offer another option for patients that need single-dose regimens and an alternative to ATV/r based second-line. However, caution is still needed since the long-term effectiveness of DTG after failure of first-line has not been evaluated and no head to head trials with ATV/r have been conducted to establish its superiority as a second-line.^26^ Therefore the durability stated here may not hold for this newer second-line in the region.

The study findings need to be interpreted with some limitations. since this was a retrospective cohort, it carries the inherent disadvantages of such a design, for example, some variables were incomplete at switch. Also, some exploratory variables, for example, employment and education level, and history of alcohol use were not tested since these are not collected in routine clinical practice. However, these socioeconomic factors affect adherence and hence viral rebound.^35^ Also, this was a single site cohort and there may be differences in practice in other centres in the region. The study examines viral rebound and no drug resistance tests were available to confirm drug resistance, and thus the study may underestimate durability. On the other hand, this retrospective population cohort is the largest and longest single centre study to examine the durability of second-line ART and to assess the clinical and demographic factors associated with viral rebound in SSA. The study uses a stringent viral load cut-off for a viral rebound and demonstrates that such cut-off could better identify those at risk of viral virological failure. The study examined several variables associated with viral rebounds. Also it being an observational study using routine data, it offers a picture of the true durability of second-line therapy in the real-world setting. Lastly, many countries have adopted the WHO guidelines, and our findings are generalisable to these HIV programs.

In conclusion, durability of second-line therapy after failure on first-line is fair, but the incidence and probability of rebound is high after eight years. Young adults, patients with a high viral load at switch, and patients switched to boosted ATV/r are more likely to experience a viral rebound. Thus, HIV clinics and programs should offer differentiated care to these risk groups with emphasis on close monitoring and enhanced adherence to maximise the durability of second-line treatment and minimise the unnecessary switch to more expensive and complicated third-line regimens.

## Data Availability

All data produced in the present study are available upon reasonable request to the authors

## Acknowledgement

We are grateful to the data management team that supported the abstraction of data and the clinical team for the feedback provided.

## Contribution of the of the authors

LI designed the study design and data analysis plan

MJ and LI analysis the data

LI and MJ draft the manuscript

CB supervised the design, data analysis, writing of the manuscript and gave critical comments on the draft manuscript.

## Declaration of interest

L.I has no conflict of interest to declare

M.J has no conflict interest to declare

CB is partly supported by the Fogarty International Centre, National Institute of Health (grant# 2D43TW009771-06 “HIV and co-infections in Uganda”).

## Data sharing agreement

The data collected including individual participant data and a data dictionary defining each field in the set will be made available to others. Such data will be provided as de-identified participant data. The study protocol and statistical analysis plan will also be available on request after publication. This data and materials will be accessed strictly by emailing the corresponding author. However, some access criteria will be applied and this will be determined by the institution’s research committee.

## Notes

### Competing Interest Statement

The authors have declared no competing interest.

### Funding Statement

This study did not receive any funding

