## Supplementary material for "Durability of second-line anti-retroviral therapy in a resource-limited setting: an 11-year analytical cohort": Table S1,Table S2,Figure S1

**Table of contents**

**Table S1: Number of viral rebound disaggregated by viral rebound cut-off and patient the characteristics……………………………………………………………………………………...................................2**

**Table S2: Table S2 Baseline factors associated with viral rebound analysed with death and lost to follow-up as competing risks using sub distribution hazard model ……………………………………………3**

**Figure S1: Incidence of viral rebound (≥ 200copies/ml) and 95% C I after switching to second line ART…………………………………………………………………………………………………………………………..4**

**Checklist S1: STROBE Checklist of items that should be included in reports of cohort studies……………………………………………………………………………………………………………………….5**

**Table S1: Number of viral rebound disaggregated by viral rebound cut-off and patient the characteristics.**

|  | **≥200 copies/ml ^§^** | | | **≥1000 copies/ml ^‡^** | | |
| --- | --- | --- | --- | --- | --- | --- |
| **Variable** | **No rebound N=746(%)** | **Viral rebound N=355 (%)** | **P -value** | **No rebound N=834(%)** | **Viral rebound N=267 (%)** | **P -value** |
| **Sex** |  |  | **0∙028** |  |  | **0∙144** |
| Male | 252 (64) | 144(34) |  | 290 (73) | 106 (27) |  |
| Female | 494 (70) | 211(30) |  | 544 (77) | 161 (23) |  |
| Age |  |  | 0∙010 |  |  |  |
| 18-24 | 33 (49) | 35(51) |  | 42 (62) | 26 (38) | 0∙019 |
| 25-34 | 245 (68) | 114(31) |  | 272 (76) | 87 (24) |  |
| 35-44 | 311(70) | 133(30) |  | 342 (77) | 102 (23) |  |
| 44-55 | 117 (67) | 58(33) |  | 134 (77) | 41 (23) |  |
| >55 | 40 (73) | 15(27) |  | 44 (80) | 11 (20) |  |
| **Viral load at switch copies/ml** |  |  | <0∙001 |  |  | <0∙001 |
| <100,000 copies/ ml | 542 (72) | 208(28) |  | 600 (80) | 150 (20) |  |
| ≥100,000copies/ml | 204 (58) | 147(42) |  | 234 (67) | 117 (33) |  |
| **CD4 at switch (cells/ul)** |  |  | 0∙285 |  |  | 0∙520 |
| <200 | 432 (67) | 212(33) |  | 485 (75) | 159 (24) |  |
| ≥200 | 220 (71) | 92(29) |  | 244 (78) | 68 (21) |  |
| **Duration of First line (ART)** |  |  | 0∙127 |  |  | 0∙156 |
| 0-3 | 280 (65) | 149(35) |  | 313 (73) | 116 (27) |  |
| 4-5 | 211 (67) | 106(33) |  | 241 (76) | 24(76 ) |  |
| >5years | 255 (72) | 100(28) |  | 280 (79) | 75 21) |  |
| **First-line NNRTI** |  |  | 0∙011 |  |  | 0∙006 |
| **EFV** | 243(63) | 145(37) |  | 273 (70) | 115 (30) |  |
| **NVP** | 470 (71) | 190 (29) |  | 522 (79) | 138 (21) |  |
| **Other** | 33 (62) | 20 (38) |  | 39 (74) | 14 (26) |  |
| **NRTI Second line regime** |  |  | 0∙776 |  |  | 0∙058 |
| TDF | 537(68) | 255(32) |  | 606 (77) | 186 23) |  |
| AZT | 69 (70) | 29(30) |  | 80 (82) | 18 18) |  |
| Others | 140(68) | 71(34) |  | 148 (70) | 63 (30) |  |
| **Anchor PI in second line** |  |  | 0∙295 |  |  | 0∙123 |
| ATV/r | 374 (69) | 166 (31) |  | 420 (78) | 120 (22) |  |
| LPV/r | 372 (66) | 189 (34) |  | 414 (73) | 147 (26) |  |
| **Calendar Year of initiation of second line** |  |  | 0∙010 |  |  | 0∙016 |
| 2007-2010 | 161(63) | 95(37) |  | 187 (73) | 69 (27) |  |
| 2011-2014 | 348(66) | 177(33) |  | 386 (74) | 139 (26) |  |
| 2015-2017 | 237 (74) | 83(26) |  | 261 (82) | 59 1(8) |  |
| **CDC score at switch** |  |  | 0∙056 |  |  | 0∙141 |
| working | 48 (80) | 12(20) |  | 51 (85) | 9 (15) |  |
| Bedridden or ambulatory | 453(68) | 212 (32) |  | 510 (77) | 155 (23) |  |
| **Hepatitis B co-infection** |  |  | 0∙855 |  |  | 0∙676 |
| Negative | 717(68) | 342(32) |  | 717 (77) | 219 (23) |  |
| Positive | 29 (69) | 13(31) |  | 31 (74) | 11 (26) |  |
| **TB co-infection** |  |  | 0∙265 |  |  | 0∙059 |
| No | 676(68) | 214(32) |  | 758 (77) | 232 (23) |  |
| Yes | 70(63) | 41(37) |  | 76 (68) | 35 (32) |  |
| **Adherence at 2nd switch** |  |  | 0∙393 |  |  | 0∙609 |
| Good | 689 (69) | 307 (31) |  | 768 (77) | 228 (23) |  |
| Fair | 3 (50) | 3 (50) |  | 4 (67) | 2 (33) |  |
| Poor | 19 (61) | 12 (39) |  | 22( 71) | 9 (29) |  |

§ Among patients that rebounded (≥200 copies/ml) median viral load was 2855 (IQR 521-37000)

‡ Among those that rebounded (≥1000 copies/ml) the median viral load 11320 (IQR 2646-79056)

P-value compares viral load cut off by variable category calculated using Х^2^

**Table S2 Baseline factors associated with viral rebound analysed with death and lost to follow-up as competing risks using sub distribution hazard model**

|  | **Unadjusted**  **SHR (95%CI)** | **P-value** | **Adjusted**  **SHR (95%CI)** | **P-value** |
| --- | --- | --- | --- | --- |
| **Sex** |  |  |  |  |
| Male | 1 |  | 1 |  |
| Female | 0∙86(0∙69-1∙06) | 0∙154 | 0∙85(0∙68- 1∙05) | 0∙135 |
| **Age group (years)** |  |  |  |  |
| 18-24 | 2∙53(1∙40- 4∙60) | 0∙002 | 2∙50(1∙34- 4∙65 ) | 0∙004 |
| 25-34 | 1∙12(0∙67- 1∙87) | 0∙672 | 1∙23(0∙71- 2∙14) | 0∙466 |
| 35-44 | 1∙01(0∙61-1∙69) | 0∙965 | 1∙06(0∙61- 1∙83) | 0∙832 |
| 44-55 | 1∙18(0∙68- 2∙05) | 0∙555 | 1∙20(0∙67- 2∙15 ) | 0∙541 |
| >55 | 1 |  | 1 |  |
| **Viral load at switch copies/ml** |  |  |  |  |
| <100,000 | 1 |  | 1 |  |
| ≥100,000 | 1∙37 (1∙11-1∙69) | 0∙003 | 1∙55(1∙24- 1∙94) | <0∙001 |
| **PI used in second-line** |  |  |  |  |
| LPV based | 1 |  | 1 |  |
| ATV based | 2∙00 (1∙58-2∙54) | <0∙001 | 1∙44(1∙10-1∙90) | 0∙008 |
| **Calendar Year of initiation of second line** |  |  |  |  |
| 2007-2010 | 1 |  | 1 |  |
| 2011-2014 | 2∙27 (1∙71-3∙01) | <0∙001 | 1∙89(1∙37- 2∙59) | <0∙001 |
| 2015-2017 | 4∙72 (3∙23-6∙88) | <0∙001 | 3∙95(2∙57- 6∙07) | <0∙001 |

**Figure S1: Incidence of viral rebound (≥ 200copies/ml) and 95% C I after switching to secondline ART**

| **Number at risk** |  |  |  |  |  |  |  |  |  |  |  |
| --- | --- | --- | --- | --- | --- | --- | --- | --- | --- | --- | --- |
| **Viral rebound** | 1082 | 960 | 759 | 591 | 456 | 343 | 252 | 154 | 97 | 49 | 15 |
| **Person Years** | 72 | 40 | 36 | 29 | 25 | 21 | 18 | 12 | 6 | 5 | 3 |

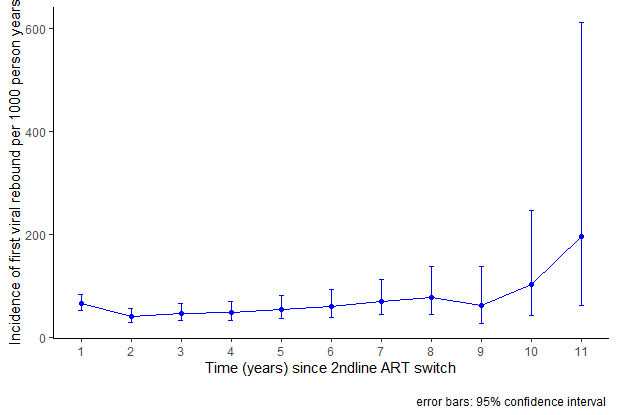

**Checklist 1: STROBE Checklist of items that should be included in reports of *cohort studies***

**Item**

**No	Recommendation**

**Title and abstract**	1

**Introduction**

Background/rationale

Objectives

2

3

Explain the scientific background and rationale for the investigation being reported

State specific objectives, including any prespecified hypotheses

(a) Indicate the study’s design with a commonly used term in the title or the abstract
(b) Provide in the abstract an informative and balanced summary of what was done and what was found

**Methods** Study Design Setting

Participants

Present key elements of study design early in the paper

Describe the setting, locations, and relevant dates, including periods of recruitment, exposure, follow-up, and data collection

4

5

(a) Give the eligibility criteria, and the sources and methods of selection of participants. Describe methods of follow-up

6

(b) For matched studies, give matching criteria and number of exposed and unexposed

Variables	7	Clearly define all outcomes, exposures, predictors, potential confounders, and effect modifiers. Give diagnostic criteria, if applicable

Data sources/ measurement

8*	For each variable of interest, give sources of data and details of methods of assessment (measurement). Describe comparability of assessment methods if there is more than one group

Bias	9	Describe any efforts to address potential sources of bias

Study size	10	Explain how the study size was arrived at

Quantitative variables	11	Explain how quantitative variables were handled in the analyses. If applicable,

describe which groupings were chosen and why

Statistical methods	12

**Results**

Participants	13*

Descriptive data	14*

(a) Describe all statistical methods, including those used to control for confounding
(b) Describe any methods used to examine subgroups and interactions
(c) Explain how missing data were addressed
(d) If applicable, explain how loss to follow-up was addressed
(e) Describe any sensitivity analyses
(a) Report numbers of individuals at each stage of study—eg numbers potentially eligible, examined for eligibility, confirmed eligible, included in the study, completing follow-up, and analysed
(b) Give reasons for non-participation at each stage
(c) Consider use of a flow diagram
(a) Give characteristics of study participants (eg demographic, clinical, social) and information on exposures and potential confounders
(b) Indicate number of participants with missing data for each variable of interest
(c) Summarise follow-up time (eg, average and total amount)

Outcome data	15*	Report numbers of outcome events or summary measures over time

Main results	16

(a) Give unadjusted estimates and, if applicable, confounder-adjusted estimates and their precision (eg, 95% confidence interval). Make clear which confounders were adjusted for and why they were included
(b) Report category boundaries when continuous variables were categorized
(c) If relevant, consider translating estimates of relative risk into absolute risk for a meaningful time period

Other analyses	17	Report other analyses done—eg analyses of subgroups and interactions, and

sensitivity analyses

**Discussion**

Key results	18	Summarise key results with reference to study objectives

Limitations	19	Discuss limitations of the study, taking into account sources of potential

bias or Discuss both direction and magnitude of any potential bias

imprecision

Interpretation	20	Give a cautious overall interpretation of results considering objectives,

limitations, multiplicity of analyses, results from similar studies, and other

relevant evidence

Generalisability	21	Discuss the generalisability (external validity) of the study results

**Other information**

Funding	22	Give the source of funding and the role of the funders for the present study and, if

applicable, for the original study on which the present article is based

*Give information separately for exposed and unexposed groups.

**Note:** An Explanation and Elaboration article discusses each checklist item and gives methodological background and published examples of transparent reporting. The STROBE checklist is best used in conjunction with this article (freely available on the Web sites of PLoS Medicine at [http://www.plosmedicine.org/,](http://www.plosmedicine.org/) Annals of Internal Medicine at [http://www.annals.org/,](http://www.annals.org/) and Epidemiology at [http://www.epidem.com/).](http://www.epidem.com/)) Information on the STROBE Initiative is available at [http://www.strobe-statement.org.](http://www.strobe-statement.org/)
